# Factors associated with viral Load re-suppression after enhanced adherence counseling among recipients of care with an Initial high viral load result in RISE-supported facilities in five States in Nigeria

**DOI:** 10.1101/2024.01.16.24301356

**Authors:** Gbenga Benjamin Obasa, Mukhtar Ijaiya, Ejike Okwor, Babafemi Dare, Franklin Emerenini, Prince Anyanwu, Adewale Akinjeji, Kate Brickson, Jennifer Zech, Yemisi Ogundare, Emmanuel Atuma, Molly Strachan, Ruby Fayorsey, Kelly Curran

## Abstract

This study explores factors influencing viral re-suppression among 1,607 recipients of HIV care in Nigeria who completed Enhanced Adherence Counseling (EAC) between July 2021 and June 2022. The analysis, part of the Reaching Impact, Saturation, and Epidemic Control (RISE) project, utilized routine program data from 132 health facilities. Results showed a 91% re-suppression rate post-EAC, with age and ART regimen type identified as significant factors. Adolescents (10-19 years) had a higher likelihood of re-suppression, while ROC on second-line regimens exhibited lower resuppression rates. These findings inform programmatic decisions to enhance HIV treatment outcomes in Nigeria.

## Background

The World Health Organization (WHO) recommends monitoring Viral load (VL) outcomes as the gold standard to measure the effectiveness of antiretroviral therapy (ART) for recipients of HIV care (ROC) [1,2]. In 2021, WHO revised its HIV treatment monitoring algorithm to assist ROC in reaching viral suppression and maintaining an undetectable VL with three categories of VL level: unsuppressed (>=1000 copies/mL), suppressed (detected but <1000 copies/mL) and undetectable (VL not detected by test used)[3]. VL is considered unsuppressed when >=1000 copies/ml for ROC undergoing treatment for more than six months. WHO recommends VL testing for ROCs on treatment for more than six months and then routinely every 12 months [1,4]. However, there are reports of low VL suppression rates among ROC living in low- and middle-income countries [5,6]. Poor adherence to HIV treatment is the most common reason for viral non-suppression and can potentially increase the risk of drug resistance, HIV treatment failure, and onward HIV transmission [7,8].

The WHO recommends Enhanced Adherence Counseling (EAC) for ROC with unsuppressed VL (>=1000 copies/ml) after six months of ART [2]. The EAC sessions lasting between 3–6 months are designed to improve adherence, identify possible barriers to treatment, and resolve treatment difficulties [5]. Studies have shown that informing ROC about the risks of an unsuppressed VL result can improve treatment adherence [9]. This will reduce the chances of developing resistance mutations, treatment failure, or disease progression to advanced HIV disease [10]. The combination of EAC with routine VL has improved outcomes among people on ART [10–12]. A first-line treatment failure is established when VL suppression is not attained after EAC sessions. Subsequently, switching to second-line treatment is considered as different ART classes are associated with varying adherence thresholds needed to achieve viral suppression and avoid resistance mutations [13].

Nigeria has the second largest HIV epidemic in the world, with a total of 1.9 million (1.4%) persons (15-49 living with HIV as reported by the Nigeria HIV/AIDS Indicator and Impact Survey (NAIIS) [14–16]. In 2020, there were around 1.8 million HIV/AIDs ROC, and about 67% of all people with HIV knew their status, but only 62% were on HIV treatment, of which 44% were estimated to be virally suppressed [16]. This VL suppression is sub-optimal compared to the UNAIDS third 95 target [15,16]. Studies have suggested several factors associated with VL non-suppression, such as poor medication adherence, suboptimal drug selection, and drug resistance [3,5,9]. Therefore, the need to assess the effectiveness of EAC in routine program implementation and determine factors associated with poor VL suppression is essential.

This analysis was conducted as part of the Reaching Impact, Saturation, and Epidemic Control (RISE) project, a five-year global project funded by the US President’s Emergency Plan for AIDS Relief (PEPFAR) through the US Agency for International Development (USAID). We analyzed factors associated with viral re-suppression for ROC who enrolled and completed all EAC sessions between July 2021 and June 2022. We analyzed the demographic and clinical characteristics of ROC with unsuppressed VL (>= 1000 copies/ml) who had completed EAC. This analysis will inform ongoing programmatic and clinical decision-making and improve health outcomes for ROC in Nigeria.

## Methodology

### Study design

A descriptive study using routinely collected service delivery data of ROC on ART in 132 RISE-supported health facilities from July 2021 to June 2022.

### Program Setting and Description

The RISE project has been implemented in 132 facilities across 56 local government areas within five states in Nigeria, beginning in 2019: Adamawa, Akwa Ibom, Cross River, Niger, and Taraba. According to NAIIS data, Akwa Ibom and Rivers have the highest number of HIV/AIDs ROC in Nigeria [15,17]. Akwa Ibom and Cross River states are located in the country’s South-South region and have a prevalence rate of 4.8% and 1.8%, respectively [15]. In the northeast, Adamawa and Taraba have HIV prevalence rates of 2.6 and 1.1 %, respectively, while Niger in the north-central zone has a prevalence of 0.9% [15].

RISE provides EAC for virally unsuppressed ROC enrolled in ART services at supported facilities in alignment with WHO guidelines. EAC is also offered to ROC who receive drugs outside the facility at the community level. Children and adolescents (<15 years) with an unsuppressed VL are also enrolled in the EAC program, with their caregivers attending the EAC sessions to assist with treatment adherence and VL suppression. According to WHO and Nigerian national guidelines, ROC with high VL results requires a repeat test after at least three EAC sessions. Recipients of care with VL results >=1000 copies/ml are enrolled into the EAC and closely monitored frequently with a minimum of 10 contacts monthly for at least three months, which can be extended to a maximum of 6 months to assess adherence level, after which a VL test is carried out post-EAC. The EAC session is designed to help ROC identify the barriers they face in taking their treatment and formulate an adherence plan. Counselors help ROC identify their goals and provide emotional support to help them reach them. After completion of the EAC sessions, if the VL result is <1000 copies/ml post-EAC, the ROC remains on the same ART regimen. If the VL result is >=1000 copies/ml after completion of the EAC sessions, the ROC is switched to a second-line ART regimen.

### Program data

De-identified client-level data from the Lafiya Management Information System (LAMIS), a web-based electronic medical system, was used to identify HIV ROC on ART with VL >= 1000 copies per ml. Client information included in the data set were sex, date of birth, ART start date, days of ART refill, last drug pickup date, ART regimen type, date of VL testing, WHO clinical staging at last visit, tuberculosis (TB) status at last visit, date of commencement of EAC, EAC enrollment, date of last EAC Session and repeat VL result after completion of EAC. Data cleaning was carried out, for instance, by converting the birth date to age and then breaking it down into age-group categories. The variable “duration on ARTV” was derived from the difference between the start date of ART and the date of last drug pickup. Our outcome variable (VL result post-EAC) was then encoded as a binary classification, which indicated whether the patient was unsuppressed (VL >= 1000 copies/ml) or resuppressed (VL detected < 1000 copies/ml).

### Study population

The analysis was conducted using data collected as part of the routine implementation of the RISE project in Nigeria. Data were collected from 132 health facilities in five states: Adamawa, Akwa Ibom, Cross River, Taraba, and Niger.

#### Inclusion criteria

All ROC enrolled in HIV care through the RISE project who have received ART for at least six months have VL results >=1000 copies/ml and who started and completed EAC sessions (at least three sessions) between July 2021 and June 2022.

#### Exclusion criteria

All ROC with no VL result, VL sample collection date, or ROC whose VL result was greater than one year were excluded. Also, ROC with data outside the selected period with no record of EAC session and documented post-EAC VL results were excluded.

### Data analysis

Data were analyzed using SAS On-Demand version 9.04. The primary outcome of the analysis was VL suppression (<1000 copies/ml) after the completed EAC session. The analysis variables were sex, age, enrollment settings, clinical staging, TB co-infection, ARV regimen line, and duration on ART. Descriptive analyses were carried out to show demographic and clinical characteristics of ROC with unsuppressed VL (>=1000 copies/ml), suppressed but detectable (>=50 to <1000 copies/ml), and undetectable VL (<50 copies/ml). In addition, Pearson’s Chi-Square was used to compare demographics, clinical characteristics, and VL re-suppression with a p-value of 0.05. Also, the Log binomial regression model was used to report crude and adjusted risk ratios with 95% Confidence Intervals (95% CI) to determine the association between clinical characteristics and VL re-suppression (<1000 copies/ml) on repeat testing, post-EAC.

### Ethical Statement

This retrospective analysis was conducted with a firm commitment to ethical principles and guidelines. The medical records were accessed for research purposes on 26th, October 2022. It is crucial to highlight that rigorous de-identification measures were implemented, ensuring the privacy and confidentiality of individual participants throughout the study. The research team did not have access to information that could identify individual participants during any phase of the research, and all data were anonymized before analysis.

The de-identification process involved the removal of any personally identifiable information and careful management of indirect identifiers that could potentially lead to participant identification. After completing the data analysis, archived samples, and records were securely stored.

### Ethical Approval

The study received ethical approval from Johns Hopkins School of Public Health Institutional Review Board IRB#24824 “Secondary Data Analysis of HIV Prevention, Care and Treatment Service Delivery Programs in Africa, which explicitly reviewed and approved the de-identification procedures to safeguard participant confidentiality.

## Results

A total of 127,198 ROC were on ART at RISE-supported Health Facilities, and 1607 (1.3%) completed EAC between July 2021 and June 2022 and had post-EAC VL results (Table 1). Among these participants, 1,054 (66%) were female. The majority, 1,410 (88%), were enrolled in facility settings, and 1,506 (88%) were on a first-line ART regimen at their last visit. Most individuals (95%) were classified as WHO clinical-stage 1 at baseline, and 1,557 (97%) were free from TB co-infection. ROC aged 40 and above accounted for 37% (589), while children < 10 years accounted for 7% (114) of ROC included in this analysis. Approximately 22% (361) of the ROC included in this analysis had been on ART for two years or less. The median time taken to complete all recommended EAC sessions was 12 weeks. Over 90% of participants completed all recommended EAC sessions within three months. Following the completion of EAC sessions, the median time for collection of a repeat VL test was 8 weeks (Table 1). About 32% of the participants had a post-EAC VL test done after 8 weeks.

**Table 1.** Demographic and clinical characteristics of Recipients of Care (ROC) with unsuppressed VL (>1000 copies/ml of blood) who completed Enhanced Adherence Counselling (EAC).

| Characteristics | N | (%) |
| --- | --- | --- |
| Total | 1607 |  |
| **Gender** |  |  |
| Female | 1054 | 65.59 |
| Male | 553 | 34.41 |
| **Age group** |  |  |
| 0-9 years | 114 | 7.09 |
| 10-19 years | 171 | 10.64 |
| 20-29 years | 226 | 14.06 |
| 30-39 years | 507 | 31.55 |
| >=40 years | 589 | 36.65 |
| **Enrollment Settings** |  |  |
| Community | 197 | 12.26 |
| Facility | 1410 | 87.74 |
| **Clinical Staging** |  |  |
| Stage I | 1537 | 95.64 |
| Stage II | 62 | 3.86 |
| Stage III | 8 | 0.50 |
| **TB Co-infection** |  |  |
| No | 1557 | 96.89 |
| Yes | 50 | 3.11 |
| **ART Regimen Line** |  |  |
| First line | 1506 | 93.71 |
| Second Line | 101 | 6.29 |
| **Duration on ARV** |  |  |
| 0-2 years | 361 | 22.46 |
| 3-5 years | 550 | 34.23 |
| 6-10 years | 463 | 28.81 |
| >10 years | 233 | 14.50 |
| **Time to complete EAC sessions** |  |  |
| Median (IQR) | 12 (0-9 weeks) |  |
| <= 12 weeks | 1516 | 94.34 |
| 13–24 weeks | 90 | 5.60 |
| ≥25 weeks | 1 | 0.06 |
| **Time from EAC session completion to repeat VL** |  |  |
| Median (IQR) | 8 (4-8 weeks) |  |
| <2 weeks | 161 | 10.13 |
| 2-4 weeks | 140 | 8.81 |
| 4-8 weeks | 515 | 32.39 |
| 8-12 weeks | 243 | 15.28 |
| ≥12 weeks | 531 | 33.40 |

*DTG-based regimens are the recommended first-line ARV drugs*
*Protease inhibitor-based regimens are the recommended second-line ARV drugs*

After the completion of EAC, a total of 1,454 individuals (91%) achieved a suppressed VL (<1000 copies/ml) (Table 2). Among them, 336 (23%) had VL >=50 - < 1000 copies/ml, and 1,118 (76%) had an undetectable VL. Females comprised 66% of those who achieved VL re-suppression after EAC. The age group of 40 years and above had the highest proportion of individuals with re-suppressed VL, totaling 542 (37%), compared to other age groups. In facility settings, 87% of ROC achieved VL re-suppression, totaling 1,267 individuals, while in community settings, 13% achieved re-suppression, comprising 187 individuals. The majority of individuals with WHO clinical-stage 1 (91%) attained VL re-suppression after EAC. Additionally, 91% of ROC without TB co-infection achieved suppressed VL after EAC (p<0.001). Furthermore, 1,408 (93%) ROC on the first-line ART regimen achieved VL re-suppression, compared to ROC on second-line ART 46 (45%) (p<0.001). ROC on treatment for more than 3-5 years had a higher proportion of VL re-suppression than those on other durations of antiretroviral therapy (p<0.031). Table 2 further details ROC characteristics and VL re-suppression in the RISE program.

**Table 2:**
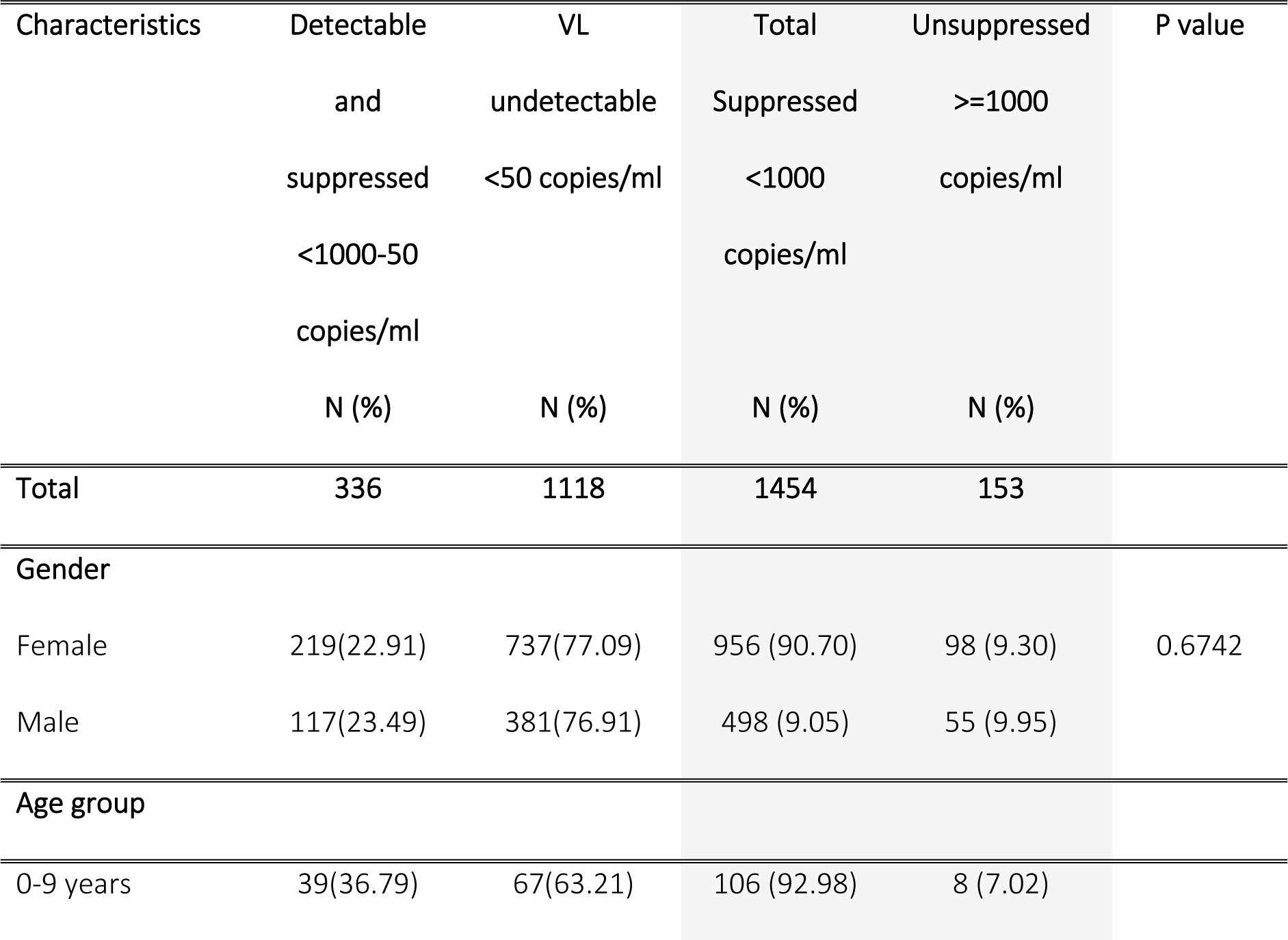
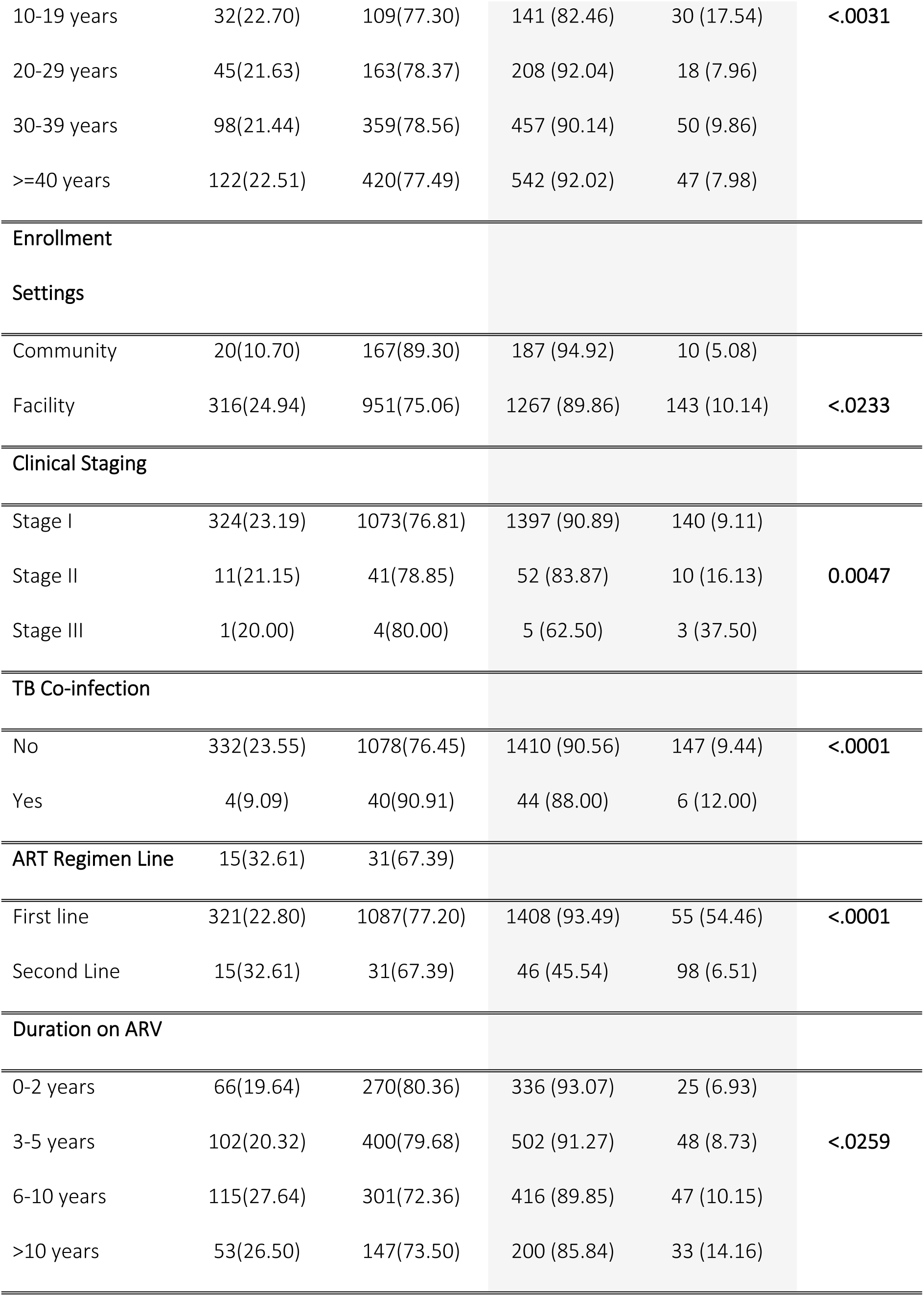
Results showing the relationship between baseline characteristics and VL resuppression

### Factors Associated with VL Re-suppression

In the unadjusted model analysis, only age, enrollment setting, ART regimen line, and ART duration were significantly associated with VL re-suppression (Table 3). ROC in the 10-19 years age band were 13% more likely to be re-suppressed compared to the 0-9 years (CRR: 1.13; 95% CI 1.04 to 1.23). Similarly, ROC in the 20 -29 years (CRR: 1.02; 95% CI 0.96 to 1.08), 30-39 years (CRR: 1.04; 95% CI 0.98 to 1.11) and >=40 years (CRR: 1.11; 95% CI 0.96 to 1.07) age bands were more likely to be re-suppressed; however, these were not statistically significant. ROC enrolled in the facility were 5.6% more likely to be resuppressed post-EAC than those enrolled in community settings (CRR: 1.06; 95% CI 1.02 to 1.10). Conversely, ROC on second-line ART regimens were 49% less likely to be re-suppressed when compared to ROC who were on first-line ART regimens at the time of analysis (CRR: 0.49; 95% CI 0.39 to 0.60). Our analysis found that the likelihood of re-suppression post-EAC increased with a longer time on treatment: 3-5 years (CRR: 1.02; 95% CI 0.98 to 1.60); 6-10 years (CRR: 1.04; 95% CI 0.99 to 1.08); and >10 years (CRR: 1.08; 95% CI 1.02 to 1.15) respectively. ROC who had been on ART treatment for more than 10 years had a statistically significant 8% higher likelihood of re-suppression than ROC who had been on ART treatment for no more than 2 years.

**Table 3.**
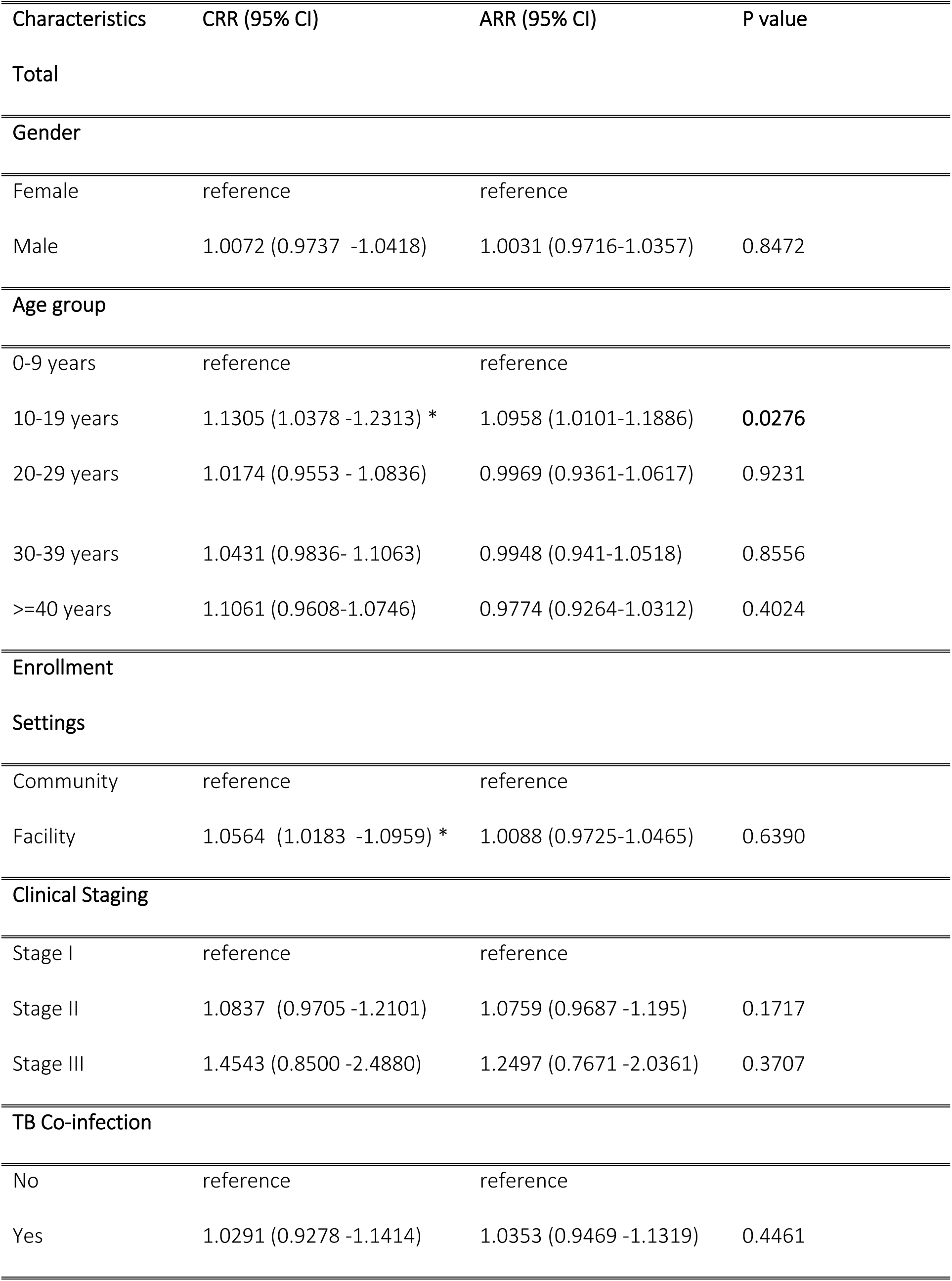
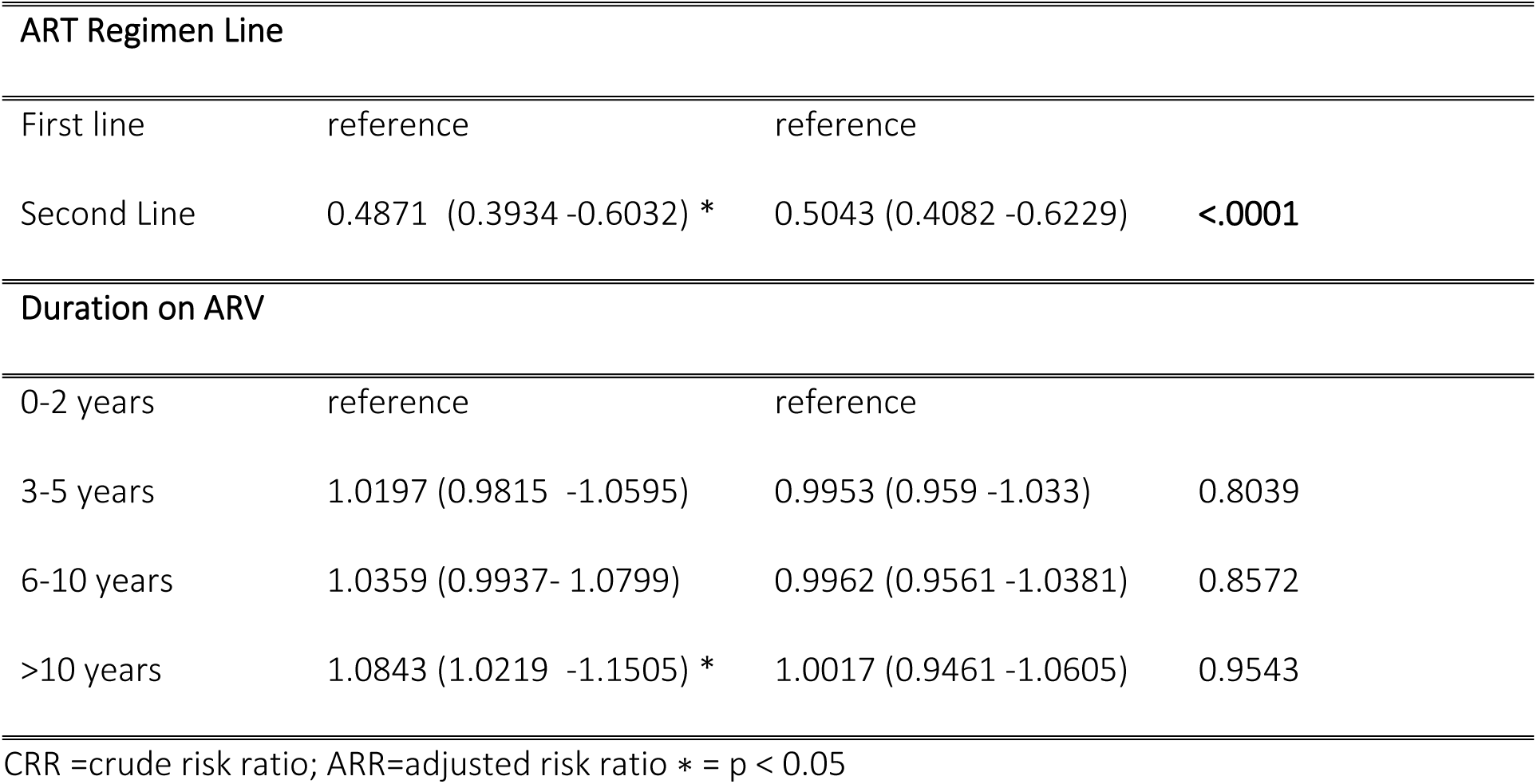
Demographic and clinical characteristics associated with resuppression of VL after completion of enhanced adherence counseling

Our adjusted model found age group and ART regimen line as the two factors significantly associated with viral re-suppression (Table 3). ROC in the 10-19 years age band were 10% more likely to be re-suppressed compared to the 0-9 years (ARR: 1.10; 95% CI 1.01 to 1.19), which was slightly lower than the probability from our bivariate analysis. While all other age bands had a reduced likelihood of re-suppression, this was not statistically significant. Notably, there was a 50% reduced likelihood of viral re-suppression among ROC on second-line regimens compared to ROC on first-line regimens (ARR: 0.50; 95% CI 0.41 to 0.62).

## Discussion

Our study explored the factors associated with VL re-suppression among 1,607 ROCs who completed EAC in RISE-supported facilities across five states in Nigeria. After completing EAC, 91% of participants achieved a suppressed VL post-EAC, with 23% having VL between >= 50 and < 1000 copies/ml and 76% having an undetectable VL. Factors associated with VL re-suppression were age and ART regimen line. Our rate of re-suppression is similar to results from other studies that showed rates of re-suppression of 90% -95% after EAC [9,13,18]. Re-suppression rates depend on EAC’s type, intensity, and documentation [13,19]. Compared to findings from other similar studies, our 91% re-suppression rate was the highest.

The demographic and clinical characteristics of the ROC in this study provided further insights into the factors associated with VL re-suppression. Age was found to be associated with VL re-suppression, with ROC aged 10-19 years significantly more likely to be re-suppressed post-EAC. This finding suggests that this age band may be associated with better post-EAC adherence and treatment outcomes. Our results may be explained by the immediate enrollment on Operation Triple Zero (OTZ), intensive follow-up of such children and adolescents, and caregiver involvement in care by the RISE team. Unlike our findings around age, other studies have found that adolescents were less likely to be re-suppressed post-EAC [9,24]. Similarly, previous studies have found that children and adolescents had comparatively lower suppression rates, with a higher probability of viral suppression among older adolescents [25,26]. Alongside poor adherence, this noted sub-optimal suppression among children and adolescents has been attributed to challenges around access to health facilities, satisfaction with healthcare services, and psychosocial factors, including stigma, school-related activities, caregiver status, knowledge, and availability [27–30].

The overall proportion of ROC with VL re-suppression did not differ between ROC enrolled in health facilities compared to those in the community. Community ART service delivery has been noted to improve ART uptake, retention, and viral suppression [31–33]. However, a study assessing facility and community refill strategies after community enrollment has noted no difference in viral suppression [34]. Moreover, the provision of comprehensive HIV care services in community settings where ROC may have relatively better access to counseling, support, and regular monitoring is effective in increasing HIV treatment coverage, and an optimal combination of both community and facility approaches will be vital in improving treatment outcomes [35].

Similar to the enrollment settings above, ROC who had been on ART for over 10 years were significantly more likely to be re-suppressed from our bivariate analysis but not in our multivariate analysis. VL suppression has been generally associated with increasing duration on treatment and adherence [26,36–38]. However, a study conducted in Ethiopia on VL among ROC post-EAC noted that ROCs on ART treatment between 3-5 years were less likely to be re-suppressed, unlike ours [18].

ART regimen was a significant predictor in our bivariate and multivariate analyses. ROC on second-line ART regimens were less likely to be resuppressed. This could be due to the majority 1,376 (91%) of ROC being on DTG-based first-line regimens per the Nigeria National Treatment Guidelines [39]. DTG-based regimens have been proven to be highly efficacious in achieving virological suppression in ROC [38,40,41]. In addition, the prior viral unsuppressed results may have been due to non-adherence, and with improved adherence following the EAC sessions, we would expect comparatively better viral re-suppression.

Furthermore, with the introduction and transition of ROC to DTG-based regimens, ROC who had been on second-line regimens may have never benefitted from the new DTG-based regimens or may have been transitioned back to the new DTG-based regimens. However, two studies on virological outcomes post-EAC in Nigeria and Zimbabwe have noted a higher likelihood of viral re-suppression among ROC on second-line ART regimens [2,21]. This has been attributed to possible lower resistance to second-line drugs and ROC’s “last chance” perception of medically responsive therapy [21]. Moreover, unsuppressed VL post-EAC has been associated with prolonged first-line ART regimen treatment and drug resistance [13].

The strength of this study is the use of routine program data, which provides valuable insights into the real-world effectiveness of EAC in Nigeria. However, there are several limitations to consider. First, the data used was cross-sectional, so causality cannot be inferred from the findings. Second, the analysis relied on routine program data, which may be subject to missing or incomplete information. Additionally, the analysis was limited to only those variables that are routinely collected from patients during visits and available in the EMR. We did not measure other potential vital factors that may be associated with VL re-suppression, such as socioeconomic factors, social support, or mental health status.

In conclusion, this study highlights the effectiveness of EAC in promoting VL suppression in routine program settings, with 91% re-suppression. The findings suggest that EAC can improve adherence to antiretroviral therapy and achieve the UNAIDS global target of 95% VL suppression. The study also identified essential factors associated with VL re-suppression, including age and ART regimen type. These findings have implications for programmatic and clinical decision-making and highlight the need for tailored interventions.

## Data Availability

The datasets used and analyzed during the current study are available from the corresponding author upon reasonable request. This analysis was carried out using data from Reaching Impact, Saturation, and Epidemic (RISE) Control program funded by the U.S. President's Emergency Plan for AIDS Relief (PEPFAR) through the United States Agency for International Development (USAID)

## References

1. World Health Organization. The use of antiretroviral drugs for treating and preventing hiv infection. 2016.

2. Bvochora T, Satyanarayana S, Takarinda KC, Bara H, Chonzi P, Komtenza B, et al. Enhanced adherence counselling and viral load suppression in HIV seropositive patients with an initial high viral load in Harare, Zimbabwe: Operational issues. PLoS ONE. 2019 Feb;14(2).

3. The Joint United Nations Programme on HIV/AIDS (UNAIDS). Understanding Fast-Track: Accelerating action to end the AIDS epidemic by 2030. 2015.

4. World Health Organization. GUIDELINE ON WHEN TO START ANTIRETROVIRAL THERAPY AND ON PRE-EXPOSURE PROPHYLAXIS FOR HIV. 2015.

5. Bulage L, Ssewanyana I, Nankabirwa V, Nsubuga F, Kihembo C, Pande G, et al. Factors Associated with Virological Non-suppression among HIV-Positive Patients on Antiretroviral Therapy in Uganda, August 2014-July 2015. BMC Infectious Diseases. 2017 May;17(1).

6. Yiltok E, Agada C, Zoakah R, Malau A, Tanyishi D, Ejeliogu E, et al. Clinical profile and viral load suppression among HIV positive adolescents attending a tertiary hospital in North Central Nigeria. Journal of Medicine in the Tropics. 2020;22(2):133.

7. Bonner K, Mezochow A, Roberts T, Ford N, Cohn J. Viral load monitoring as a tool to reinforce adherence: A systematic review. Journal of Acquired Immune Deficiency Syndromes. 2013 Sep;64(1):74–8.

8. Charurat M, Oyegunle M, Benjamin R, Habib A, Eze E, Ele P, et al. Patient Retention and Adherence to Antiretrovirals in a Large Antiretroviral Therapy Program in Nigeria: A Longitudinal Analysis for Risk Factors. Myer L, editor. PLoS ONE. 2010 May;5(5):e10584.

9. Jobanputra K, Parker LA, Azih C, Okello V, Maphalala G, Kershberger B, et al. Factors associated with virological failure and suppression after enhanced adherence counselling, in children, adolescents and adults on antiretroviral therapy for HIV in Swaziland. PLoS ONE. 2015 Feb;10(2).

10. Labhardt ND, Bader J, Lejone TI, Ringera I, Hobbins MA, Fritz C, et al. Should viral load thresholds be lowered?: Revisiting the WHO definition for virologic failure in patients on antiretroviral therapy in resource-limited settings. Medicine. 2016 Jul;95(28):e3985.

11. WHO. world health Organisation. 2016. The Use of Antiretroviral Drugs for treating And Preventing HIV infection guidelines HIV/AIDS Programme.

12. Jobanputra K, Parker LA, Azih C, Okello V, Maphalala G, Jouquet G, et al. Impact and programmatic implications of routine viral load monitoring in swaziland. Journal of Acquired Immune Deficiency Syndromes. 2014 Sep;67(1):45–51.

13. Ford N, Orrell C, Shubber Z, Apollo T, Vojnov L. HIV viral resuppression following an elevated viral load: a systematic review and meta-analysis. Journal of the International AIDS Society. 2019 Nov;22(11).

14. National Agency for the Control of AIDS (NACA). NATIONAL HIV AND AIDS STRATEGIC FRAMEWORK 2021-2025. 2022.

15. Federal Ministry of Health N. Nigeria HIV/AIDS Indicator and Impact Survey (NAIIS) 2018: Technical Report. Abuja; 2019.

16. Monitoring GA. Country progress report -Nigeria. 2020;

17. Negedu-Momoh OR, Balogun O, Dafa I, Etuk A, Oladele EA, Adedokun O, et al. Estimating HIV incidence in the Akwa Ibom AIDS indicator survey (AKAIS), Nigeria using the limiting antigen avidity recency assay. Journal of the International AIDS Society. 2021 Feb;24(2):e25669.

18. Diress G, Dagne S, Alemnew B, Adane S, Addisu A. Viral Load Suppression after Enhanced Adherence Counseling and Its Predictors among High Viral Load HIV Seropositive People in North Wollo Zone Public Hospitals, Northeast Ethiopia, 2019: Retrospective Cohort Study. AIDS research and treatment. 2020;2020:8909232.

19. Etoori D, Ciglenecki I, Ndlangamandla M, Edwards CG, Jobanputra K, Pasipamire M, et al. Successes and challenges in optimizing the viral load cascade to improve antiretroviral therapy adherence and rationalize second-line switches in Swaziland. Journal of the International AIDS Society. 2018 Oct;21(10):e25194.

20. Ali MW, Musa MS. The effect of mobile phone utilization for enhanced adherence counselling intervention among persons with HIV. AIDS Care. 2023 Feb 13;1–9.

21. Awolude OA, Olaniyi O, Moradeyo M, Abiolu J. Virologic Outcomes Following Enhanced Adherence Counselling among Treatment Experienced HIV Positive Patients at University College Hospital, Ibadan, Nigeria. International STD Research & Reviews. 2021 Mar 2;10(1 SE-Original Research Article):53–65.

22. Laxmeshwar C, Acharya S, Das M, Keskar P, Pazare A, Ingole N, et al. Routine viral load monitoring and enhanced adherence counselling at a public ART centre in Mumbai, India. PLOS ONE. 2020 May 5;15(5):e0232576.

23. Ukwueze LN, Ifeanyichukwu C, Ifeanyi C, Iroegbu A, Aribike J, Ikeneche NF. Evaluation of Enhanced Adherence Counselling among Virally Unsuppressed HIV-Infected Adults on Antiretroviral Therapy in Suburban and Metropolitan parts of Delta State , Nigeria. 2021;5(04).

24. Mhlanga TT, Jacobs BKM, Decroo T, Govere E, Bara H, Chonzi P, et al. Virological outcomes and risk factors for non-suppression for routine and repeat viral load testing after enhanced adherence counselling during viral load testing scale-up in Zimbabwe: analytic cross-sectional study using laboratory data from 2014 to 2018. AIDS Research and Therapy. 2022;19(1):34.

25. Nasuuna E, Kigozi J, Babirye L, Muganzi A, Sewankambo NK, Nakanjako D. Low HIV viral suppression rates following the intensive adherence counseling (IAC) program for children and adolescents with viral failure in public health facilities in Uganda. BMC Public Health. 2018;18(1):1048.

26. Chhim K, Mburu G, Tuot S, Sopha R, Khol V, Chhoun P, et al. Factors associated with viral non-suppression among adolescents living with HIV in Cambodia: a cross-sectional study. AIDS research and therapy. 2018 Nov;15(1):20.

27. Montalto GJ, Sawe FK, Miruka A, Maswai J, Kiptoo I, Aoko A, et al. Diagnosis disclosure to adolescents living with HIV in rural Kenya improves antiretroviral therapy adherence and immunologic outcomes: A retrospective cohort study. PLOS ONE. 2017 Oct 9;12(10):e0183180.

28. Eticha T, Berhane L. Caregiver-reported adherence to antiretroviral therapy among HIV infected children in Mekelle, Ethiopia. BMC Pediatrics. 2014;14(1):114.

29. Bikaako-Kajura W, Luyirika E, Purcell DW, Downing J, Kaharuza F, Mermin J, et al. Disclosure of HIV status and adherence to daily drug regimens among HIV-infected children in Uganda. AIDS and behavior. 2006 Jul;10(4 Suppl):S85–93.

30. Gichane MW, Sullivan KA, Shayo AM, Mmbaga BT, O’ Donnell K, Cunningham CK, et al. Caregiver role in HIV medication adherence among HIV-infected orphans in Tanzania. AIDS care. 2018 Jun;30(6):701–5.

31. Labhardt ND, Ringera I, Lejone TI, Klimkait T, Muhairwe J, Amstutz A, et al. Effect of Offering Same-Day ART vs Usual Health Facility Referral During Home-Based HIV Testing on Linkage to Care and Viral Suppression Among Adults With HIV in Lesotho: The CASCADE Randomized Clinical Trial. JAMA. 2018 Mar;319(11):1103–12.

32. Eshun-Wilson I, Awotiwon AA, Germann A, Amankwaa SA, Ford N, Schwartz S, et al. Effects of community-based antiretroviral therapy initiation models on HIV treatment outcomes: A systematic review and meta-analysis. PLOS Medicine. 2021 May 28;18(5):e1003646.

33. Barnabas R V, Szpiro AA, van Rooyen H, Asiimwe S, Pillay D, Ware NC, et al. Community-based antiretroviral therapy versus standard clinic-based services for HIV in South Africa and Uganda (DO ART): a randomised trial. The Lancet Global health. 2020 Oct;8(10):e1305–15.

34. Amstutz A, Lejone TI, Khesa L, Kopo M, Kao M, Bresser M, et al. Offering ART Refill Through Community Health Workers Versus Clinic-Based Follow-Up After Home-Based Same-Day ART Initiation: The VIBRA Cluster-Randomised Clinical Trial. 2021;

35. Oladele EA, Badejo OA, Obanubi C, Okechukwu EF, James E, Owhonda G, et al. Bridging the HIV treatment gap in Nigeria : examining community antiretroviral treatment models. 2018;

36. Tomescu S, Crompton T, Adebayo J, Akpan F, Dauda DS, Allen Z, et al. Factors associated with viral load non-suppression in people living with HIV on ART in Nigeria: cross-sectional analysis from 2017 to 2021. BMJ open. 2023 May;13(5):e065950.

37. van Liere GAFS, Lilian R, Dunlop J, Tait C, Rees K, Mabitsi M, et al. High rate of loss to follow-up and virological non-suppression in HIV-infected children on antiretroviral therapy highlights the need to improve quality of care in South Africa. Epidemiology and infection. 2021 Mar;149:e88.

38. Mehari EA, Muche EA, Gonete KA. Virological Suppression and Its Associated Factors of Dolutegravir Based Regimen in a Resource-Limited Setting: An Observational Retrospective Study in Ethiopia. HIV/AIDS (Auckland, NZ). 2021;13:709–17.

39. Federal Ministry of Health Nigeria. National Guidelines for HIV Prevention, Treatment and Care. 2020.

40. Abudiore O, Amamilo I, Campbell J, Eigege W, Harwell J, Conroy J, et al. High acceptability and viral suppression rate for first-Line patients on a dolutegravir-based regimen: An early adopter study in Nigeria. PLOS ONE. 2023 May 17;18(5):e0284767.

41. Nabitaka VM, Nawaggi P, Campbell J, Conroy J, Harwell J, Magambo K, et al. High acceptability and viral suppression of patients on Dolutegravir-based first-line regimens in pilot sites in Uganda: A mixed-methods prospective cohort study. PLOS ONE. 2020 May 27;15(5):e0232419.

